# Perceived barriers and facilitators to geriatric trauma risk assessment: Instrument development and validation

**DOI:** 10.1101/2023.06.29.23292067

**Authors:** Oluwaseun Adeyemi, Sanjit Konda, Corita Grudzen, Charles DiMaggio, Garrett Esper, Erin Rogers, Keith Goldfeld, Saul Blecker, Joshua Chodosh

## Abstract

**Background:** In the fast-paced and high-stress environment of the ED, conducting a thorough and efficient risk assessment is may be associated with various challenges such as time constraints and competing priorities. The aim of this study is to develop and validate a survey instrument that will comprehensively assesses emergency provider and nurses perceived barriers and facilitators to geriatric trauma risk assessment.

**Methods:** We designed two six-item survey that each assesses the barriers and facilitators to geriatric trauma risk assessment using the American College of Surgeons geriatric trauma management guidelines. Each item in the survey has a quantitative section, answered on a binary scale, and a qualitative open ended responses. Nine content experts performed content validation of the items in the scale and we computed Cohen’s Kappa, and item and scale content validity indices (CVIs).

**Results:** Most of the experts were male (56%), and non-Hispanic Whites (44%). A third of the content experts are MDs. Of the six items in the perceived barriers scale, five items were retained. The Cohen’s Kappa value across the five items ranged from 0.4 to 0.9 and the item and scale CVIs for the five items were 0.76 each. Of the six items in the perceived facilitators to geriatric trauma risk assessment, all six items were retained. The Cohen’s Kappa value across the six items was 1.00 and the item and scale CVIs for the six items were 1.00 each.

**Conclusion:** We presents an instrument that can assess the perceived barriers and facilitators to geriatric trauma risk assessment experienced by emergency providers and nurses.

## Introduction

Geriatric trauma risk assessment is a vital process that aims to identify and evaluate the specific risks and vulnerabilities faced by older adults with traumatic injuries. While traumatic injuries can affect individuals of all age groups, older adults are particularly vulnerable due to a combination of physiological, cognitive, and psychosocial factors.^1-3^ Physiologically, older adults have reduced bone density leading to higher risk of fractures, thinner and more fragile skin leading to incrased risk of bruises and lacerations, and a decline in muscle strength and balance leading to increased risk of falls.^4,5^ Age-related cognitive changes, such as memory impairment or cognitive decline, can further impact an older adult’s ability to perceive and respond to potential hazards or risky situations and increase the risk of injuries.^6,7^ Psychosocial factors, such as social isolation, unsafe living conditions, and substance use can increase the risk of injuries and hinder access to prompt medical care.^8,9^

Risk assessment in the emergency department (ED) involves a systematic evaluation of the patient’s condition, focusing on identifying potential life-threatening injuries, immediate interventions needed, and the overall risk profile of the patient.^10,11^ For geriatric trauma patients, particular attention is paid to their risk factors, such as frailty^12-16^ and pre-existing medical conditions,^17,18^ which can influence injury patterns and the severity of trauma. Cognitive function and baseline functional status are also assessed to understand their capacity to participate in their own care and rehabilitation.^19^ The information gathered during the geriatric trauma risk assessment process guides decision-making regarding further diagnostic tests, consultations with specialists, and the development of an appropriate care plan. It aids in determining the need for surgical interventions, pain management strategies, and preventive measures to minimize the risk of complications and optimize outcomes.

In the fast-paced and high-stress environment of the ED, conducting a thorough and efficient risk assessment is crucial to identify the severity of injuries, prioritize interventions, and ensure appropriate and timely care delivery. Yet achieving a thorough and efficient risk assessment may be associated with various challenges such as time constraints and competing priorities may limit the depth and thoroughness of the risk assessment process. Additionally, the lack of knowledge of the unique characteristics and complexities of the geriatric trauma population may serve as a barrier in creating a high index of suspicion for the synergistic effect of underlying comorbidities, polypharmacy, frailty, functional limitations, and cognitive impairement. Assessing the barriers as well as the facilitators to effective geriatric trauma risk assessment may inform on the need for educational intervention that will improve emergency providers and nurses healthcare service delivery. However, there is no validated instrument in the extant geriatric trauma literature that measures, quantitatively or qualitatively, emergency providers and nurses perceived barriers to geriatric trauma risk assessment. The aim of this study is to develop and validate a survey instrument that will comprehensively assess emergency provider and nurses perceived barriers and facilitators to geriatric trauma risk assessment.

## Methods

### Study Design and Population

We employed a purposive sampling technique to select survey instrument experts to assess the content validity of the items in the perceived barriers and facilitators to geriatric trauma risk assessment. The study population were content and instrument experts. This validation study is part of the project aimed at developing geriatric trauma triage and risk assessment (Institutional Review Board: s15-00371).

### Inclusion and Exclusion Criteria

Our content and instrument experts must be either healthcare providers or clinical research scientists. Each expert must have spent at least a year in clinical practice or in clinical research. Clinicians without research experience were excluded. Also, clinical scientist without knowledge of basic emergency care were excluded.

### Scale Development

The items in the perceived barriers and facilitators to geriatric trauma risk assessment were extracted from the American College of Surgeons Geriatric Trauma Quality Improvement survey.^20^ We identified six domains of risk assessment: depression, substance use, high-risk medications, frailty, functional limitations from activities of daily living, and comprehensive geriatric care. For the barrier domain, we created six items that assessed perceived barriers to geriatric trauma risk assessment. Each item has a quantitative and qualitative sections. The quantitative aspect is measured on a dichotomous scale (yes or no) and the qualitative aspect is an open-ended question to an affirmative response to the leading question. Similarly, for the facilitator domain, we created six items that assessed perceived enablers to geriatric trauma risk assessment. Each item has quantitative and qualitative parts. The quantitative aspect is measured on a dichotomous scale (yes or no) and the qualitative aspect is an open-ended question to an affirmative response to the leading question.

### Analytical Plan

We reported the demographic and occupation characteristics of the content and instrument experts. To assess the content validity of the items related to perceived barriers and facilitators in geriatric trauma risk assessment, we computed the content validity index (CVI). ^21^ The instrument experts evaluated the relevance of each item in the barrier and facilitator scales using a four-level Likert-type scale (1 - irrelevant; 2 - unable to assess relevance without revision; 3 - relevant but needs minor alteration; 4 - extremely relevant). The qualitative section included prompts such as “if yes, please explain.” We converted the four-level scale into a binary scale, considering items coded as 1 to be relevant (either relevant or relevant with minor alterations), while all other responses were coded as irrelevant (coded as 0). For each item, we calculated the item content validity index (I-CVI) as the mean score, and we assessed the experts’ agreement on the relevance of each item. To determine the agreement, we calculated Cohen’s Kappa using the formula: 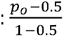, where *p*_*0*_ was the observed relevant proportion.^22^

Items with a Cohen’s kappa value of 0.2 or greater were retained.^21^ Furthermore, we computed the scale content validity index (S-CVI) in two steps: firstly, by determining the proportion of experts who agreed on the relevance of items in the perceived barriers and facilitators scales, and secondly, by calculating the average of these proportions to obtain the S-CVI.^21^ The survey was distributed using Research Electronic Data Capture (REDCap), ^23^ and the data were analyzed using IBM Statistical Package for Social Sciences (SPSS) version 28. ^24^

## Results

### Demographic and Occupational Characteristics

Nine content and instrument experts examined the perceived barriers and facilitators survey (Table 1). The mean (SD) age of the experts was 34.0 (8.3) years. The experts were mostly males (56%) and non-Hispanic White (44%). Thirty-three percent of the experts are MDs, 55% had master degrees, and 33% had doctoral degrees. The median years of practice was six years.

**Table 1:** Demographic and occupational characteristics of the content experts

| Variables | Frequency (N=9) |
| --- | --- |
| Age (Mean (SD)) | 34.0 (8.3) |
| Sex (n(%)) |  |
| Male | 5 (55.6) |
| Female | 4 (44.4) |
| Race/Ethnicity (n(%)) |  |
| Non-Hispanic White | 4 (44.4) |
| Non-Hispanic Black | 1 (11.1) |
| Multi-race | 2 (22.2) |
| Other Races | 2 (22.2) |
| Educational Qualification* (n(%)) |  |
| Master | 5 (55.6) |
| PhD | 3 (33.3) |
| M.D. | 3 (33.3) |
| Years of practice** (Median (Q1, Q3)) | 6.0 (3.0, 12.0) |

\*Multiple options, proportion exceeds 100%; Years of practice refers to clinical practice

### Content Validity: Perceived barriers and facilitators to geriatric trauma risk assessment

Among the six items in the perceived barriers to geriatric trauma risk assessment, one item had Cohen’s Kappa value of less than 0.2, and it was removed (Table 2). Of the remaining five items, the Cohen’s Kappa value ranged from 0.33 to 0.78. The item and scale CVIs for the five items were each 0.76. Among the six items in the perceived facilitators to geriatric trauma risk assessment, all the items perfect agreement and Cohen’s Kappa value of 1.0. (Table 3). The item and scale CVIs for the six items were each 1.00.

**Table 2:**
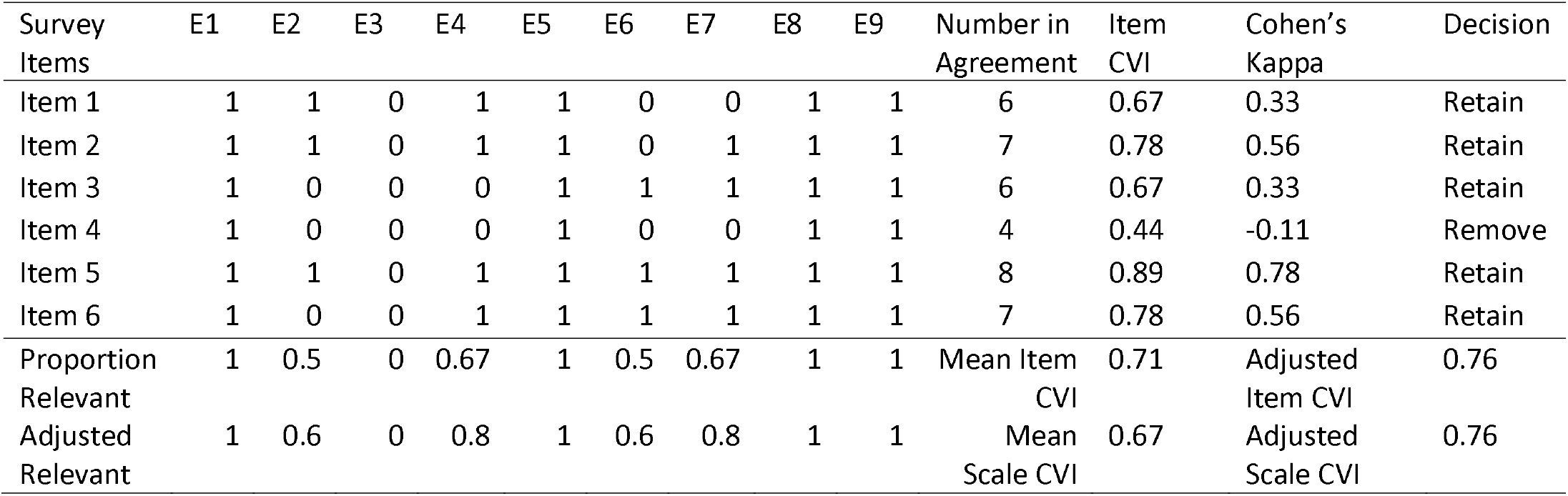
Summary of content validity index of the items on the perceived barriers to geriatric trauma assessment

| Survey Items | E1 | E2 | E3 | E4 | E5 | E6 | E7 | E8 | E9 | Number in Agreement | Item CVI | Cohen's Kappa | Decision |
| --- | --- | --- | --- | --- | --- | --- | --- | --- | --- | --- | --- | --- | --- |
| Item 1 | 1 | 1 | 0 | 1 | 1 | 0 | 0 | 1 | 1 | 6 | 0.67 | 0.33 | Retain |
| Item 2 | 1 | 1 | 0 | 1 | 1 | 0 | 1 | 1 | 1 | 7 | 0.78 | 0.56 | Retain |
| Item 3 | 1 | 0 | 0 | 0 | 1 | 1 | 1 | 1 | 1 | 6 | 0.67 | 0.33 | Retain |
| Item 4 | 1 | 0 | 0 | 0 | 1 | 0 | 0 | 1 | 1 | 4 | 0.44 | -0.11 | Remove |
| Item 5 | 1 | 1 | 0 | 1 | 1 | 1 | 1 | 1 | 1 | 8 | 0.89 | 0.78 | Retain |
| Item 6 | 1 | 0 | 0 | 1 | 1 | 1 | 1 | 1 | 1 | 7 | 0.78 | 0.56 | Retain |
| Proportion Relevant | 1 | 0.5 | 0 | 0.67 | 1 | 0.5 | 0.67 | 1 | 1 | Mean Item CVI | 0.71 | Adjusted Item CVI | 0.76 |
| Adjusted Relevant | 1 | 0.6 | 0 | 0.8 | 1 | 0.6 | 0.8 | 1 | 1 | Mean Scale CVI | 0.67 | Adjusted Scale CVI | 0.76 |

**Table 3:** Summary of content validity index of the items on the perceived facilitators to geriatric trauma assessment

| Survey Items | E1 | E2 | E3 | E4 | E5 | E6 | E7 | E8 | E9 | Number in Agreement | Item CVI | Kappa | Decision |
| --- | --- | --- | --- | --- | --- | --- | --- | --- | --- | --- | --- | --- | --- |
| Item 1 | 1 | 1 | 1 | 1 | 1 | 1 | 1 | 1 | 1 | 9 | 1.00 | 1.00 | Retain |
| Item 2 | 1 | 1 | 1 | 1 | 1 | 1 | 1 | 1 | 1 | 9 | 1.00 | 1.00 | Retain |
| Item 3 | 1 | 1 | 1 | 1 | 1 | 1 | 1 | 1 | 1 | 9 | 1.00 | 1.00 | Retain |
| Item 4 | 1 | 1 | 1 | 1 | 1 | 1 | 1 | 1 | 1 | 9 | 1.00 | 1.00 | Retain |
| Item 5 | 1 | 1 | 1 | 1 | 1 | 1 | 1 | 1 | 1 | 9 | 1.00 | 1.00 | Retain |
| Item 6 | 1 | 1 | 1 | 1 | 1 | 1 | 1 | 1 | 1 | 9 | 1.00 | 1.00 | Retain |
| Proportion Relevant | 1 | 1 | 1 | 1 | 1 | 1 | 1 | 1 | 1 | Mean Item CVI | 1.00 | Adjusted Item CVI | 1.00 |
| Adjusted Relevant | 1 | 1 | 1 | 1 | 1 | 1 | 1 | 1 | 1 | Mean Scale CVI | 1.00 | Adjusted Scale CVI | 1.00 |

## Discussion

We present a content-validated instrument suitable for assessing the perceived barriers and facilitators to geriatric trauma risk assessment experienced by emergency providers and nurses. Designed using the American College of Surgeons’s geriatric trauma guidelines, this instrument can serve as a tool to assess the individual and institutional factors associated with adherence and its lack, to geriatric trauma risk assessment. Of the original six items in both perceived barriers domain, one item was excluded. All the items in the perceived facilitators were retained.

The item ‘when you meet a geriatric trauma patient, will you encounter barriers in screening for medication use that can affect initial evaluation and care?’ was excluded from the perceived barriers domain. One possible reason for this exclusion could be the growing recognition of the significance of medication reconciliation for geriatric trauma patients.^25-28^ Medication errors are considered never events and serious reportable events,^29^ highlighting the need for a safe and effective process to document and communicate a patient’s medications throughout their care journey. Healthcare providers and institutions are required to prioritize and enforce policies that ensure medication reconciliation is implemented during care transitions, as mandated by the U.S. Joint Commission.^30^ Although medication reconciliation may present challenges,^31^ the potential risks associated with medication errors that could harm patients outweigh any barriers. Importantly, it is worth noting that this item was not excluded from the perceived facilitators domain.

The content validation process employed in this study has certain limitations. Our group of content experts consisted of both clinicians and non-clinicians with knowledge of routine clinical practice. The clinical knowledge may have introduced a bias towards including certain types of questions. This bias could explain the varying Cohen’s kappa scores observed in the perceived barriers subdomain. Additionally, the selection of experts was conducted through purposive sampling, and the lead researcher was aware of their identities. However, the lack of blinding is unlikely to have had a differential impact on the survey items since the independent experts were unaware of each other’s responses. Despite these limitations, the study has notable strengths. It is the first known study to develop an instrument capable of quantitatively and qualitatively capturing the perceived barriers and facilitators to geriatric trauma risk assessment among emergency providers and nurses. As a result, this survey provides a valuable tool for evaluating and gaining deeper insights into the challenges associated with geriatric trauma risk assessment.

## Conclusion

We presents an instrument that can assess the perceived barriers and facilitators to geriatric trauma risk assessment experienced by emergency providers and nurses. By integrating open ended responses to the items in the perceived barrier and facilitator instrument, investigators can gain deeper insight into the challenges associated with geriatric trauma assessment among emergency providers.

## Data Availability

All data produced in the present study are available upon reasonable request to the authors

## Notes

### Competing Interest Statement

The authors have declared no competing interest.

### Funding Statement

This study did not receive any funding

### Author Declarations

New York University Grossman School of Medine Institutional Review Board approved this study: s15-00371

